# Retrospective meta-transcriptomic identification of severe dengue in a traveller returning from Africa to Sweden, 1990

**DOI:** 10.1101/2020.09.08.20184580

**Authors:** Kristian Alfsnes, Nina Lagerqvist, Sirkka Vene, Jon Bohlin, Jenny Verner-Carlsson, David Ekqvist, Andreas Bråve, Edward C. Holmes, Weifeng Shi, John H.-O. Pettersson

## Abstract

The first imported case of severe haemorrhagic fever in Sweden was reported in 1990. Despite extensive diagnostic study, no aetiological agent was identified. Following retrospective investigation with total RNA-sequencing of plasma and urine samples collected during between 7–36 days from onset of symptoms, we identified dengue virus 3 (DENV-3) and a human pegivirus (HPgV). We conclude that the patient most likely suffered from haemorrhagic symptoms due to a severe dengue infection.

## Background

Currently, more than 20 enveloped RNA viruses are known to cause haemorrhagic fever, largely from four groups (arenaviruses, filoviruses, bunyaviruses, and flaviviruses) [1]. These pathogens can induce symptoms ranging from asymptomatic to severe life-threatening conditions, with some haemorrhagic fevers associated with mortality rates as high as 80% [1]. However, a major complicating factor is that the initial clinical presentations commonly overlap with syndromes that have a less severe clinical course. A rapid diagnosis is therefore of major importance.

We utilized RNA-sequencing (‘meta-transcriptomics’) to investigate Sweden’s first case of imported haemorrhagic fever for which the aetiological agent was never identified. The background, initial handling and laboratory analyses, clinical data, epidemiology, and outcome have been described previously [2,3]. Briefly, a male Swedish traveller returned from a three-month visit in central and eastern Africa in January 1990, the last month of which was spent in Kenya. Five days after arriving in Sweden, the patient fell ill with febrile symptoms. On the fourth day after onset of symptoms, the patient’s condition deteriorated rapidly, whereafter he was admitted to a hospital and shortly thereafter moved to an intensive care unit. Soon after transferal to the intensive care unit, symptoms expanded to include internal haemorrhaging. During the haemorrhagic episode, which began at the start of week two and stopped at the end of the third week after symptom onset, the patient had received 65 units of blood. It is also noteworthy that the patient had an extensive febrile period, with temperatures consistently above 40°C, that lasted for approximately three weeks despite symptomatic treatment. His clinical course also included sepsis, respiratory failure and disseminated intravascular coagulation with thrombocytopenia. After 4.5 weeks, the patient’s condition improved and he was moved from the intensive care unit to a regular ward. He was discharged 2.5 months after onset of symptoms.

Initial differential diagnoses excluded leptospirosis, malaria, rickettsiosis, typhoid fever and septicaemia. Despite attempts with cell-, rodent- and primate inoculation, serological tests, blood smears and electron microscopy, no aetiological agent was identified and no definitive diagnosis was reached. However, it was suggested that the haemorrhagic fever might be due to a novel virus, potentially an undescribed filovirus as indicated by electron microscopy images taken during the first week of symptoms [2].

## Methods

We performed a retrospective meta-transcriptomic investigation using a set of separately aliquoted − 80°C stored plasma and urine samples collected between 7–18 days and 28–36 days after onset of symptoms, respectively (Table 1). Oral and written informed consent was obtained from the patient. This approach allows for a sensitive and unbiased identification and characterization of all RNA transcripts in a particular sample with the potential to identify both known and novel pathogens [4]. The methodology used herein has been described previously [4,5]. Briefly, total RNA was extracted using a combination of trizol–chloroform separation and automated magnetic bead total nucleic acid extraction using a MagLEAD system (Precision System Science Co.). Following ribosomal RNA depletion using the Ribo-Zero Gold Kit (Illumina), paired-end (100bp read-length) libraries where created with (i) the TruSeq stranded TotalRNA kit (Illumina) and sequenced on an Illumina NovaSeq 6000 instrument at the Science for Life Laboratory, Uppsala, Sweden, and (ii) Trio RNA-seq library preparation kit (NuGen) and sequenced on the BGI BGI500 (paired-end, 100bp read-length) platform at BGI, Hong Kong. Subsequently, all RNA sequence libraries were quality trimmed with Trimmomatic v.0.36 (www.usadellab.org/cms/?page=trimmomatic) and human reads were removed using bbmap (https://jgi.doe.gov/data-and-tools/bbtools/) and the human reference genome (RefSeq assembly: GCF_000001405.38). The quality-passed and human-trimmed reads were then screened with Diamond v.0.9.15.116 (https://github.com/bbuchfink/diamond) and blastn against the complete non-redundant protein (nr) and nucleotide (nt) databases, respectively.

## Results

Following inspection of the read hits to known viruses, we were able to identify sequence reads with high similarity against dengue virus (DENV) in 11 out 15 sequencing libraries. No other potential pathogens were identified, aside from human pegivirus (HPgV), formerly known as GB virus C, in five of the libraries (Table 1), which is usually associated with benign infections [6].

All reads of DENV origin were retrieved and re-screened on NCBI blast to identify the most similar reference genome. Accordingly, a complete DENV-3 genome collected 1985 in Mozambique (GenBank accession FJ882575.1) was found to exhibit the greatest sequence similarity to the DENV reads present in the patient libraries. Subsequently, all individual sequence libraries were mapped against the DENV-3 reference genome from Mozambique using bbmap, resulting in a total, following merging of read mapping files, of 768 reads (incl. duplicate reads) covering approximately 38% (4,055 of 10,562 bp) of the viral genome. Similarly, all sequence libraries were mapped against a HPgV reference genome from USA (MK291245.1) using bbmap, resulting in a total of 115 reads (incl. duplicate reads) for all HPgV positive libraries covering ca. 18% (1,641 of 9,319 bp) of the reference genome.

To place the partial DENV-3 genome in an evolutionary and epidemiological context, closely related sequences were retrieved from NCBI/GenBank and aligned using Mafft v.7.271 (https://mafft.cbrc.jp/alignment/software/) employing the E-INS-i algorithm. A maximum likelihood tree was then inferred, via IQ-TREE v.1.6.11 (http://www.iqtree.org/), using the TIM2+F+**r**_4_ model of nucleotide substitution and a Shimodaira–Hasegawa-like test to assess branch support. The resultant tree was viewed, annotated in FigTree (https://github.com/rambaut/figtree) and mid-point rooted. The DENV-3 sequence identified here shared a most recent common ancestor (88.6% bootstrap support) with the reference sequence from Mozambique, compatible with an African origin for the Swedish infection (Figure 1). The same analytical procedure was performed HPgV using the GTR+F+R6 model of nucleotide evolution. The partial HPgV sequence was found within a group of viruses of diverse geographic origins, although including east Africa (81.5% bootstrap support) (Figure 1).

The meta-transcriptomic findings were confirmed with real-time PCR, commercial ELISA (Dengue Virus IgM Capture DxSelect, Focus Diagnostics), and immunofluorescence analysis detecting dengue virus IgG antibodies [7] (Figure 1). Although negative results can never definitively rule out an infection of a different aetiological agent, all samples that tested positive for DENV with real-time PCR [8], yet tested negative for Ebola virus, Crimean-Congo Haemorrhagic Fever virus, Marburg virus, Rift Valley fever virus, and Lassa virus using PCR.

All sequence libraries generated in this project, in which human reads have been removed, have been deposited at the NCBI Short Read Archive under BioProject ID: XXXX. The partial DENV-3 consensus genome generated has been deposited in NCBI GenBank (accession number: YYYY).

**Table 1.** RNA-seq, PCR and serological results for each sample

| Sampling date | Sample ID | Material | DENV |  |  | HPgV |  |  |
| --- | --- | --- | --- | --- | --- | --- | --- | --- |
|  |  |  | RNAseq | # of reads | PCR | RNAseq | # of reads | PCR |
| 1990-01-24 | OA | Plasma | + | 57 | + | + | 53 | – |
| 1990-01-24 | OB | Plasma | + | 14 | + | – | 0 | + |
| 1990-01-30 | 1B | Plasma | + | 30 | + | + | 8 | + |
| 1990-01-30 | 1C | Plasma | + | 24 |  | + | 38 |  |
| 1990-01-31 | 2A | Plasma | + | 32 |  | + | 8 |  |
| 1990-01-31 | 2B | Plasma | + | 18 | + | + | 8 | + |
| 1990-02-01 | 3 | Plasma | + | 124 |  | – | 0 |  |
| 1990-02-02 | 4A | Plasma | + | 213 |  | – | 0 |  |
| 1990-02-02 | 4B | Plasma |  |  | + |  |  | – |
| 1990-02-04 | 5A | Plasma | + | 80 |  | – | 0 |  |
| 1990-02-04 | 5B | Plasma | + | 30 | + | – | 0 | – |
| 1990-02-04 | 5C | Plasma | + | 146 |  | – | 0 |  |
| 1990-02-14 | 6 | Urine | – | 0 | – | – | 0 |  |
| 1990-02-18 | 7 | Urine | – | 0 | – | – | 0 |  |
| 1990-02-18 | 8 | Urine | – | 0 |  | – | 0 |  |
| 1990-02-22 | 9 | Urine | – | 0 | – | – | 0 |  |

\*Onset of symptoms = 1990-01-17

**Figure 2.**
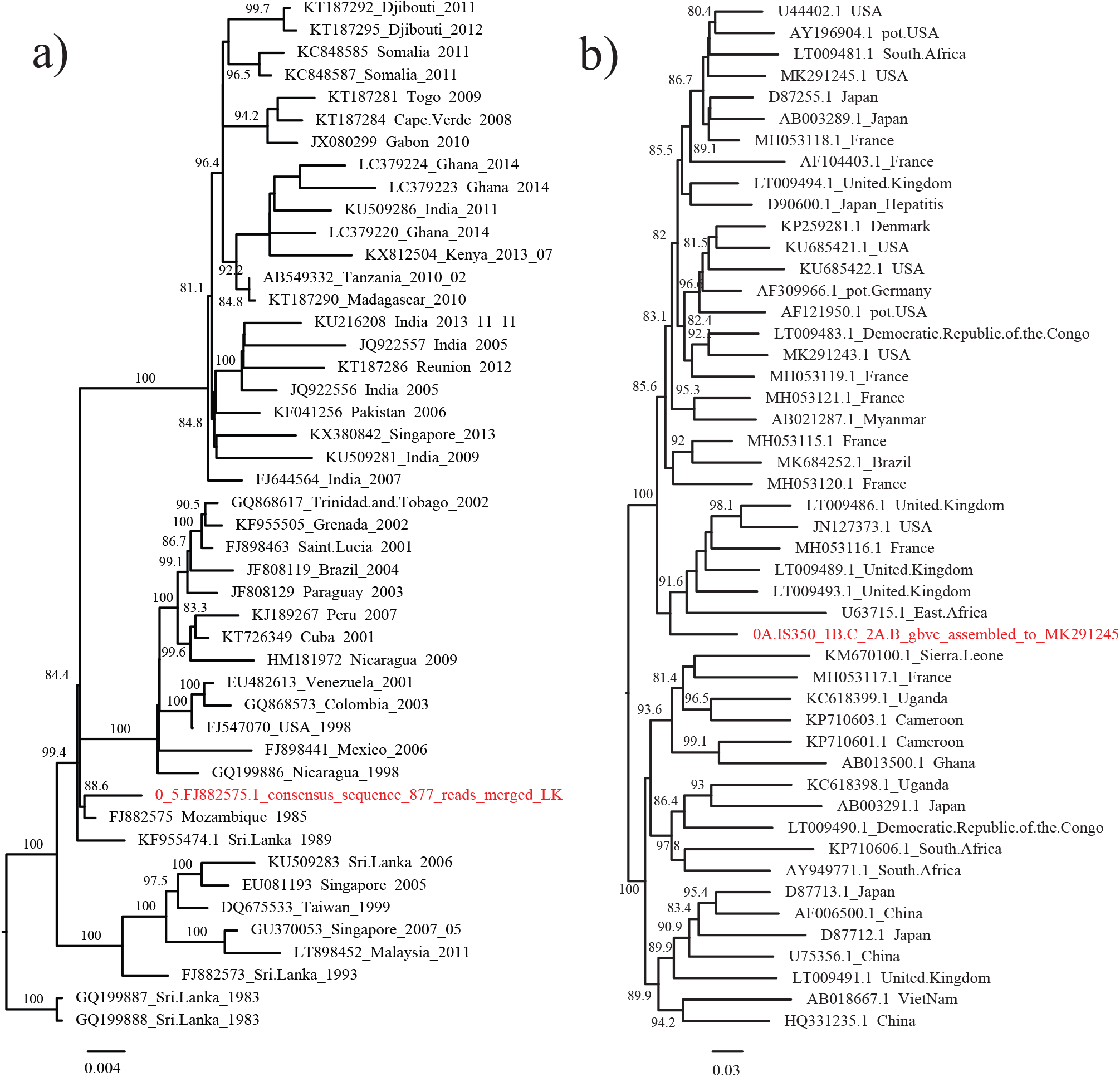
Phylogenetic trees of (a) DENV-3 and (b) HPgV, respectively, with representative sequences. The tree were mid-point rooted for visual purposes only. Numbers on branches indicate bootstrap support above 80%. Scale-bars indicates the number of nucleotide substitutions per site.

## Discussion

Using a meta-transcriptomic approach we identified DENV-3 as the likely aetiological agent of an undiagnosed severe haemorrhagic fever episode in a patient returning to Sweden following travel to central and eastern Africa in 1990.

Over the last 50 years the four serotypes of DENV have spread globally and become established in almost all regions, where previously only one serotype was present. Although DENV is likely endemic in many African countries, infections with this virus have only been reported sporadically from African countries [9], with DENV-3 first identified in Mozambique in 1984 [10]. Phylogenetic analysis shows that the DENV-3 identified here groups with the sequence from Mozambique from the 1984–1985 epidemic, compatible with an African origin (Figure 1). This adds support to the conclusion that the patient was infected with DENV-3, most likely during his final one-month stay in Kenya, a location where DENV is greatly under-sampled. A DENV-3 infection might also explain why there were no secondary cases to any of the care givers or laboratory staff at the hospital, although direct blood exposure was reported [2].

In five of the sequencing libraries (Table 1), partly confirmed with PCR, we noted the presence of HPgV, phylogenetically indicated to be of non-African origin (Figure 1). HPgV is a common virus, present in up to 5% of healthy blood-donors [6], and is most closely related to simian pegiviruses [11]. HPgV is currently not definitively associated with any disease [12], and the majority of individuals clear the infection within two years [6]. However, HPgV co-infection has been linked to increased survival rates in Ebola virus and human immunodeficiency virus patients [13,14]. As HPgV was found to be present before and during the haemorrhagic episode in the present study, we cannot draw any firm conclusions as for the potential clinical impact of this.

In conclusion, we show that total RNA-sequencing is a usable and sensitive tool to investigate undiagnosed cases caused by an infectious aetiological agent, with the potential to identify possible viral co-infections. Our study also adds support to the notion that dengue virus has most likely been present, potentially endemic, in many sub-Saharan African countries for decades, possible causing unrecognized cases of both dengue fever and more severe dengue disease. Increased surveillance efforts will aid in more accurately estimating the true burden of dengue in sub-Saharan African countries where it is clearly heavily underestimated [15].

## Data Availability

All sequence libraries generated in this project, in which human reads have been removed, are under process of being deposited at the NCBI Short Read Archive. This also applies for the partial DENV-3 and HPgV consensus genome generated which will be deposited at NCBI GenBank.

## Acknowledgement

JHOP is funded by the Swedish research council FORMAS (grant no: 2015–710). ECH is funded by an ARC Australian Laureate Fellowship (FL170100022).

## References

1. Paessler S, Walker DH. Pathogenesis of the viral hemorrhagic fevers. Annu Rev Pathol. 2013; 8:411-440.

2. Foberg U, Frydén A, Isaksson B, et al. Viral haemorrhagic fever in Sweden: experiences from management of a case. Scand J Infect Dis. 1991; 23(2): 143-151.

3. Kenyon RH, Niklasson B, Jahrling PB, et al. Virologic investigation of a case of suspected haemorrhagic fever. Res Virol. 1994; 145(6):397–406.

4. Pettersson JH-O, Piorkowski G, Mayxay M, et al. Meta-transcriptomic identification of hepatitis B virus in cerebrospinal fluid in patients with central nervous system disease. Diagn Microbiol Infect Dis. 2019; 95(4):114878.

5. Ling J, Persson Vinnersten T, Hesson JC, et al. Identification of hepatitis C virus in the common bed bug - a potential, but uncommon route for HCV infection? Emerg Microbes Infect. 2020; 9(1):1429–1431.

6. Bhattarai N, Stapleton JT. GB virus C: the good boy virus? Trends Microbiol. 2012; 20(3):124–130.

7. Vene S, Mangiafico J, Niklasson B. Indirect immunofluorescence for serological diagnosis of dengue virus infections in Swedish patients. Clin Diagn Virol. 1995; 4(1):43–50.

8. Alm E, Lesko B, Lindegren G, et al. Universal single-probe RT-PCR assay for diagnosis of dengue virus infections. PLoS Negl Trop Dis. 2014; 8(12):e3416.

9. Amarasinghe A, Kuritsky JN, Letson GW, Margolis HS. Dengue virus infection in Africa. Emerging Infect Dis. 2011; 17(8):1349–1354.

10. Gubler DJ, Sather GE, Kuno G, Cabral JR. Dengue 3 virus transmission in Africa. Am J Trop Med Hyg. 1986; 35(6):1280–1284.

11. Porter AF, Pettersson JH-O, Chang W-S, et al. Novel hepaci- and pegi-like viruses in native Australian wildlife and non-human primates. Virus Evolution [Internet]. 2020 [cited 2020 Aug 29];. Available from: https://academic.oup.com/ve/advance-article/doi/10.1093/ve/veaa064/5894981

12. Stapleton JT, Foung S, Muerhoff AS, Bukh J, Simmonds P. The GB viruses: a review and proposed classification of GBV-A, GBV-C (HGV), and GBV-D in genus Pegivirus within the family Flaviviridae. J Gen Virol. 2011; 92(Pt 2):233-246.

13. Heringlake S, Ockenga J, Tillmann HL, et al. GB virus C/hepatitis G virus infection: a favorable prognostic factor in human immunodeficiency virus-infected patients? J Infect Dis. 1998; 177(6):1723–1726.

14. Lauck M, Bailey AL, Andersen KG, Goldberg TL, Sabeti PC, O’Connor DH. GB virus C coinfections in west African Ebola patients. J Virol. 2015; 89(4):2425–2429.

15. Bhatt S, Gething PW, Brady OJ, et al. The global distribution and burden of dengue. Nature. 2013; 496(7446):504–507.

